# Profile of red blood cell disorders among healthy schoolchildren in the low malaria-endemic region in Indonesia

**DOI:** 10.64898/2026.07.20.26358521

**Authors:** Ayodhia Pitaloka Pasaribu, Illene Nanine, Framita Ainur, Vincent Jimanto, Azkarunia Pasidiaz Hutagalung, Lydia Visita Panggalo, Devin, Olga Rasiyanti Siregar, Beby Syofiani Hasibuan, Fahmi Fahmi, Leily Trianty, Farah Novita Coutrier, R. Tedjo Sasmono, Ari Winasti Satyagraha

## Abstract

Red blood cell (RBC) disorders arose as an advantageous evolutionary response to malaria infections. In a heterozygous condition, such as in Southeast Asian Ovalocytosis (SAO), hosts are protected against severe malaria. In malaria-endemic regions, RBC disorders are presumed to be highly prevalent. Tanjung Leidong, a moderately endemic area in North Sumatra (API 1.13 in 2024), lacks comprehensive data on RBC disorder prevalence beyond G6PD deficiency. Therefore, this study aims to characterize the RBC disorders in this region as well as to characterize the anemia status in children living in Tanjung Leidong. Schoolers attending D. I. Panjaitan elementary to high school were recruited and screened for malaria by microscopy and G6PD deficiency using the STANDARD G6PD Assay. The DNA of the participants was also extracted to be genotyped for SAO, Hemoglobin E (HbE), and α-thalassemia. Exclusively, G6PD-deficient DNA samples were genotyped further to determine variants. The proportion of G6PD deficiency, SAO, HbE, α-thalassemia one-gene deletion, and two-gene deletion were 0.90%, 0.90%, 2.40%, 6.26%, and 0.30%, respectively. Anemia prevalence was approximately 14%, and RBC disorders were observed across children with normal to obese BMI. No malaria infections were detected by microscopy. The predominance of asymptomatic RBC disorders highlights that they are protective against malaria infection, although their protective role against malaria could not be directly assessed in this study. Both nutritional and genetic factors are found to contribute to anemia in this cohort. These findings underscore the importance of integrated screening strategies for RBC disorders and anemia in malaria-endemic settings.

## Introduction

Malaria has been circulating for thousands of years and imposes significant selective pressure on the human body, particularly red blood cells (RBCs), to evolve in order to protect itself [1]. The protection manifests in the form of clinical symptoms that lead to RBC disorders, though some conditions might be asymptomatic until exposed to certain agents or infections [2]. These disorders differ in how they affect the RBCs. Nine amino acid deletions in the band 3 protein causes Southeast Asian Ovalocytosis (SAO) by modifying the skeletal and membrane protein interactions of the RBC membrane, which increases the rigidity of the membrane, causing early removal in the spleen [1]. Enzymatic disorders, such as glucose-6-phosphate-dehydrogenase (G6PD) deficiency, lower enzyme stability, making the RBC more susceptible to oxidative stress [4]. Genetic mutations in the hemoglobin-coding genes lead to reduced expression of the affected globin chains, resulting in hemoglobinopathy or thalassemia. For example, point mutation in hemoglobin E (HbE) which results in imbalance production of globin chains, changes the hemoglobin structure and function as well as the RBC morphology [5]. All these effects would render the parasite difficult to infiltrate and/or to multiply in these RBCs, thus, offering protection against severe malaria. From an evolutionary standpoint, SAO has disadvantages in that, it is embryonic lethal in the homozygous state, thus, only heterozygous state can survive. Similarly, thalassemia traits could both confer protection and create clinical complications. While individuals with HbE trait alone are generally asymptomatic, the combination of HbE with β-thalassemia or sickle cell disease, can lead to severe complications including anemia, organ enlargement, bone deformities, and increased risk of infections [6]. Another example, G6PD-deficient individuals, who are normally asymptomatic, can experience acute hemolytic anemia when triggered by highly oxidative drugs. These adaptations illustrate a balance between protection from malaria and potential negative consequences [7]. Natural selection causes those with advantageous mutations to survive and pass on these mutations, compared to those that cause clinical symptoms. Over time, this selective pressure increases the frequency of protective mutations in malaria-endemic populations, even if some of these mutations have trade-offs in the form of RBC disorders [8].

Malaria holds a significant burden of mortality and morbidity to this day, with approximately 263 million cases worldwide causing about 567 thousand deaths [9]. It is a vector-borne disease caused by the *Plasmodium* protozoan and transmitted through *Anopheles sp* mosquitoes. Among them, *P. falciparum*, *P. vivax*, *P. malariae*, and *P. ovale* are pathogenic to human [10]. According to World Health Organization in 2023, in Southeast Asia, there are four million malaria cases, with Indonesia as the second highest contributor with one million cases in 2022. With that in mind, efforts in research and discoveries for malaria treatments and prevention have been made to eliminate the disease. Currently, medications used to treat malaria include 8-aminoquinoline family of drugs, such as primaquine and tafenoquine, which are potent to eliminate the liver stages of *P. vivax*. However, these drugs are highly oxidative, which could compromise G6PD-deficient patients due to their vulnerable RBCs [11]. Consequently, this highlights the importance of screening RBC disorders as it ensures treatments and improves patient outcomes, especially in endemic areas. Therefore, having accurate prevalence data in these regions is essential to strategically administer safe radical cure [12]. Additionally, certain RBC disorders can cause pathological neonatal jaundice, which is a critical concern in areas where the primary treatment of phototherapy is not always available in remote healthcare centers. Anticipating the burden of RBC disorders through prevalence data helps to plan healthcare infrastructure and clinical management [1, 13]. Another importance of mapping RBC disorders is the resulting chronic anemia that may lead to childhood stunting. Recognizing these underlying causes are important because treatment options will differ as nutritional supplementation may not address anemia caused by genetic disorders [14]. For example, iron supplements administered to anemic patients may be detrimental to thalassemia patients. Despite the recognized importance of understanding RBC disorder prevalence in malaria-endemic settings, such data remain limited at the subnational and community levels in Indonesia.

Tanjung Leidong is an area in the district of Labuhanbatu Utara, North Sumatra, where it is known to be moderate-endemic for malaria, with 704 reported cases in 2023 [15] and API of 1.13 in 2024 (based on the Ministry of Health of the Republic of Indonesia). Currently, there is no information disclosing the prevalences of RBC disorders, except for G6PD deficiency, in this area. Early detection of RBC disorders is crucial for timely intervention and prevention, as it enables appropriate nutritional and medical management [16]. Consequently, it is important to monitor these areas and develop treatment strategies tailored to the local conditions. Thus, this study aims to characterize RBC disorders in healthy schoolchildren of Tanjung Leidong while also looking at their anemia status and risk of hemolysis caused by 8-aminoquinolines.

## Materials and Methods

### Ethics statement

This study has been approved by the Research Ethics Committee of the Universitas Sumatra Utara (975/KEPK/USU/2024) and Health Research Ethics Committee of National Research and Innovation Agency (065/KE.04/SK/04/2025), stating the research has been conducted based on the principles in the Declaration of Helsinki. Written informed consent was obtained from all 334 healthy participants. Parents or guardians signed the consent form for minors.

### Study site and population

Tanjung Leidong is a sub-district in the Kualuh Leidong District (API value of 1.13) located in the northern region of Labuhanbatu Utara regency, North Sumatra (Fig 1). This cross-sectional study was conducted at the D. I. Panjaitan Foundation, which comprises of an elementary school to senior high school in the same complex, throughout August 12^th^ to September 4^th^, 2024. The total study population consisted of 473 students from the foundation, with a final sample size of 334 subjects (Fig 2). There are many ethnic groups within the study population with Batak being the dominant ethnic group followed by Javanese, Chinese, Malay, Padang, Banjar, Bugis, Nias, and Sasak. The inclusion criteria included healthy students currently attending the D. I. Panjaitan Foundation and living in the area, and who were willing to participate and signed an assent form or obtained informed consent from their parents/legal guardians. Participants who refused or had severe illnesses were excluded.

**Fig 1.**
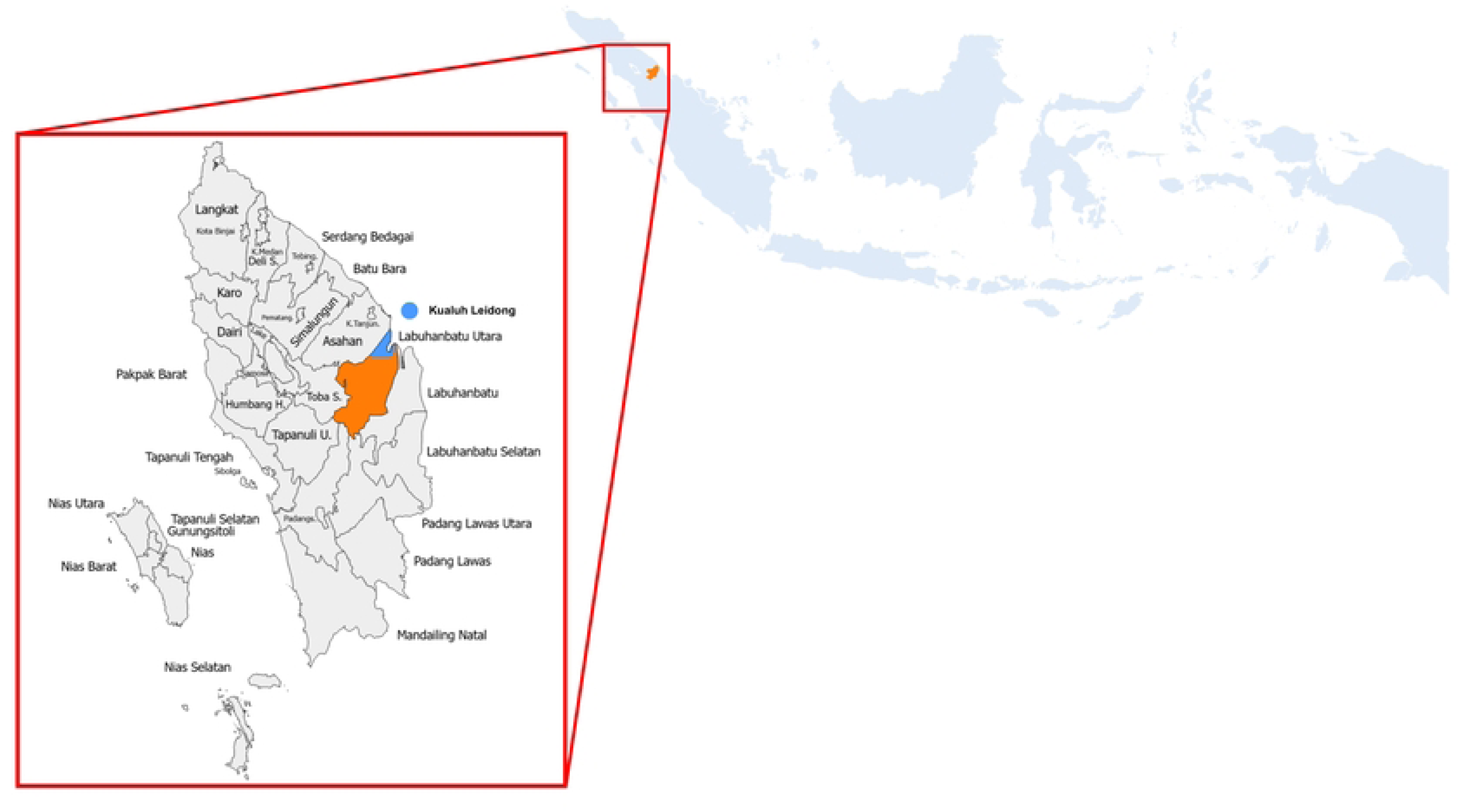
Map of Tanjung Leidong, Kualuh Leidong District, North Labuhanbatu regency.

**Fig 2.**
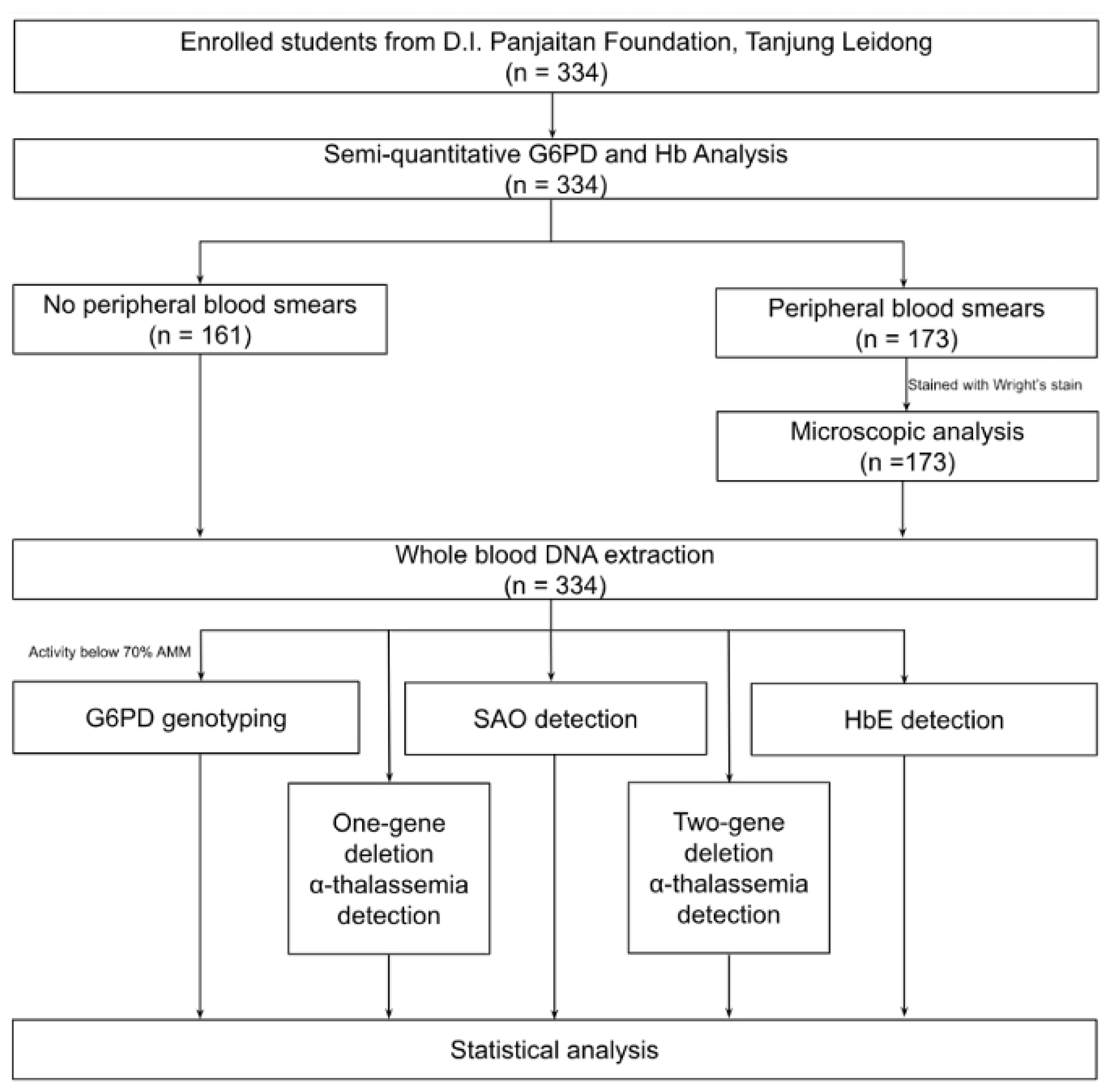
Schematic workflow of the study in Tanjung Leidong in 2024.

### Body mass index calculation

Participants’ height and weight were measured on-site. Body Mass Index (BMI) was calculated and categorized as described by World Health Organization (2025).

### Malaria identification and G6PD deficiency detection using SD Biosensor Standard G6PD test

Participants were identified for malaria through microscopy analysis according to standardized procedures [18]. Afterwards, approximately 500 μL of capillary blood were collected in EDTA-microtainer tubes. All subjects were assessed for G6PD enzyme activity using the STANDARD G6PD Test (SD Biosensor, South Korea). The test is based on a colorimetric system by detecting the innate fluorescence of nicotinamide adenine dinucleotide phosphate (NADPH) utilizing approximately 10 µL of the capillary blood to determine G6PD activity and hemoglobin level. To compare hemoglobin measurement, another 10 μL of blood was analyzed using HemoCue Hb301+ system (HemoCue AB, Sweden). Both tests were done on-site. Blood samples were kept at 4°C and transported to Jakarta for further analysis. Samples with G6PD activity below 70% of the Adjusted Male Median (AMM) were genotyped. The AMM was calculated to account for the variability of activities due to severe G6PD deficiency, and the adjusted median was set as 100% G6PD activity. Samples with activity below 30% AMM were deemed deficient. Meanwhile, intermediate level fell between the 30% and 70% AMM [19].

### G6PD variant genotyping using polymerase chain reaction and restriction fragment length polymorphism analysis

G6PD-deficient samples were selected to be genotyped. Genomic DNA from whole blood samples was extracted with the QIAamp DNA Blood Mini kit (QIAGEN, Hilden, Germany) per manufacturer’s instruction. DNA concentration and quality were measured using the Nanophotometer N60/N50 (IMPLEN, Munich, Germany). Polymerase chain reaction (PCR) and restriction fragment length polymorphism (RFLP) for G6PD variants common to Indonesian population were performed based on the protocols described by Satyagraha et al. (2016). Other variants were described also in other literature [21–25]. Samples with seemingly normal results after PCR-RFLP will be sent for Sanger Sequencing (1st BASE, Singapore) and the resulting sequences were aligned to a reference sequence from NCBI NG_009015.2 [26].

### SAO, HbE, and α-thalassemia genotyping

Extracted DNA samples were genotyped for SAO, HbE, and α-thalassemia (one-gene and two-gene deletions). List of primers used and PCR conditions for genotyping were as written in Satyagraha et al. (2016).

### Statistical analysis

This study employed the Bland-Altman plot to assess the agreement between hemoglobin levels measured by HemoCue and those by the STANDARD G6PD Test. Furthermore, the numbers of detected RBC disorders were utilized to calculate the final proportion of RBC disorders in schoolchildren of Tanjung Leidong. The mean, median, standard deviation, and range of G6PD enzymatic activities were calculated. Data were analyzed using R software.

## Results

### Clinical characteristics, demographic, and proportion of RBC disorders

A total of 334 schoolchildren were screened for this study. Table 1 lists their demographic characteristics along with the RBC disorders analysis. There was a similar distribution of females (51.2%) and males (48.8%) in this study. The mean age of participants was 13.20 (3.42) years, which ranged from 6–20 years old. Most of the ethnicity was dominated by Batak (47%), followed by Javanese (26%) and Chinese (19%). Hemoglobin levels mostly ranged from 10–15 g/dL, with an average of 13.50 (1.67) g/dL. When compared to the WHO classification for anemia based on hemoglobin, the number of females with anemia almost doubled that of males, with one male having severe anemia. About 5.6% of the children were found to have moderate anemia (Table 2). BMI analysis showed that most participants have a normal weight status. The STANDARD G6PD Test identified three males as deficient and two females as intermediate, indicating a proportion of 0.9% for G6PD deficiency. Genotyping also revealed a proportion of 0.9% for SAO, 2.4% for HbE, 6.3% for α-thalassemia one-gene deletion, and 0.3% for α-thalassemia two-gene deletion. One subject exhibited a double mutation of heterozygous SAO and homozygous α-thalassemia one-gene deletion (3.7 kb).

**Table 1.** Demographic table of schoolchildren in D. I. Panjaitan Foundation, Tanjung Leidong.

| Variable | Mean<sup>a</sup> | Total Samples<br>N = 334<sup>b</sup> |
| --- | --- | --- |
| **Sex** |  |  |
| Female |  | 171 (51.20) |
| Male |  | 163 (48.80) |
| **Age (years)** |  |  |
| < 7 | 6.0 (0.00) | 9 (2.69) |
| 7–12 | 9.9 (1.7) | 122 (36.53) |
| 13–18 | 15.4 (1.6) | 198 (59.28) |
| > 18 | 19.2 (0.4) | 5 (1.50) |
| **Ethnicity** |  |  |
| Banjar |  | 1 (0.30) |
| Batak |  | 156 (46.71) |
| Batak & Chinese |  | 1 (0.30) |
| Bugis |  | 1 (0.30) |
| Chinese |  | 63 (18.86) |
| Javanese |  | 86 (25.75) |
| Malay |  | 21 (6.29) |
| Nias |  | 1 (0.30) |
| Padang |  | 3 (0.90) |
| Sasak (Lombok) |  | 1 (0.30) |
| **Hemoglobin (g/dL)** |  |  |
| < 10 |  | 3 (0.90) |
| 10–15 |  | 280 (83.83) |
| > 15 |  | 51 (15.27) |
| **BMI Category<sup>17</sup> (kg)** |  |  |
| Underweight | 14.9 (1.5) | 46 (13.77) |
| Normal | 18.4 (2.5) | 228 (68.26) |
| Overweight | 23.7 (2.8) | 28 (8.38) |
| Obese | 29.0 (5.1) | 32 (9.58) |
| **G6PD Status and Activity** |  |  |
| Deficient (< 30%) |  | 3 (0.90) |
| Intermediate (30–70%) |  | 3 (0.90) |
| Normal (> 70%) |  | 328 (98.20) |
| **SAO** |  |  |
| Heterozygous |  | 3 (0.90) |
| Homozygous normal |  | 331 (99.10) |
| **HbE** |  |  |
| Heterozygous |  | 8 (2.40) |
| Homozygous normal |  | 326 (97.60) |
| **α-Thalassemia One-Gene Deletion** |  |  |
| Homozygous 3.7 kb deletion |  | 9 (2.69) |
| Heterozygous 3.7 kb deletion |  | 3 (0.90) |
| Heterozygous 4.2 kb deletion |  | 9 (2.69) |
| Normal |  | 313 (93.71) |
| **α-Thalassemia Two-Gene Deletion** |  |  |
| Heterozygous --<sup>SEA</sup> |  | 1 (0.30) |
| Normal |  | 333 (99.70) |

BMI: body mass index, G6PD: glucose-6-phosphate dehydrogenase, SAO: Southeast Asian ovalocytosis, HbE: hemoglobin E
<sup>a</sup>n (SD)
<sup>b</sup>n (%)

**Table 2.** Comparison of anemia status in males and females in the community.

| Variable | Female<br>(N = 171) | Male<br>(N = 163) |
| --- | --- | --- |
| **Anemia<sup>a</sup>** |  |  |
| Mild anemia |  |  |
| Children, age 5–11 years (11.0–11.4 g/dL) | 0 (0.00) | 1 (0.61) |
| Children, age 12–14 years (11.0–11.9 g/dL) | 4 (2.45) | 3 (1.84) |
| Girls, age 15–17 years (11.0–11.9 g/dL) | 7 (4.29) | 0 (0.00) |
| Boys, age 15–17 years (11.0–12.9 g/dL) | 0 (0.00) | 2 (1.23) |
| Moderate anemia (8.0–10.9 g/dL) | 12 (7.02) | 7 (4.29) |
| Severe anemia (< 8.0 g/dL) | 0 (0.00) | 1 (0.61) |
| Normal |  |  |
| Children, age 5–11 years (> 11.5 g/dL) | 43 (25.15) | 51 (31.29) |
| Children, age 12–14 years (>12.0 g/dL) | 38 (22.22) | 39 (23.93) |
| Girls, age 15–17 years (>12.0 g/dL) | 67 (39.18) | 0 (0.00) |
| Boys, age 15–17 years (>13.0 g/dL) | 0 (0.00) | 59 (36.20) |

<sup>a</sup>n (%)
Based on World Health Organization (2024)

### Proportion of G6PD deficiency

The AMM was calculated to be 8.72 U/g Hb and used to determine the cutoff thresholds. Hence, the 30% and 70% AMM were calculated respectively as 2.46 and 5.74 U/g Hb. Fig 3 illustrates the distribution of subjects across the G6PD activity values with the thresholds of 10%, 30%, and 70%. A total of six samples with activity <70% of normal G6PD were genotyped. One male subject with an intermediate G6PD activity (5.6 U g/Hb) was genotyped and no mutation was found. Genotyping results of the rest of 5 samples revealed that Vanua Lava (c.383 T > C) was the most common variant (3/5), followed by Canton (c.1376 G > T) and Viangchan (c.871 G > A). Compared to another study in the area, the most common variant was Mahidol (c.487 G > A) [27] with G6PD prevalence of 2.6%. Females with intermediate activity were 1.17% (2/169), compared to males who were deficient at about 1.84% (3/160). Between deficient and normal groups, there was a significant difference in the activity, which may result in adverse effects, on those who are deficient when treated with high dose of primaquine or tafenoquine. Despite living in a malaria-endemic area, all participants in this study were tested negative for malaria by microscopy.

**Fig 3.**
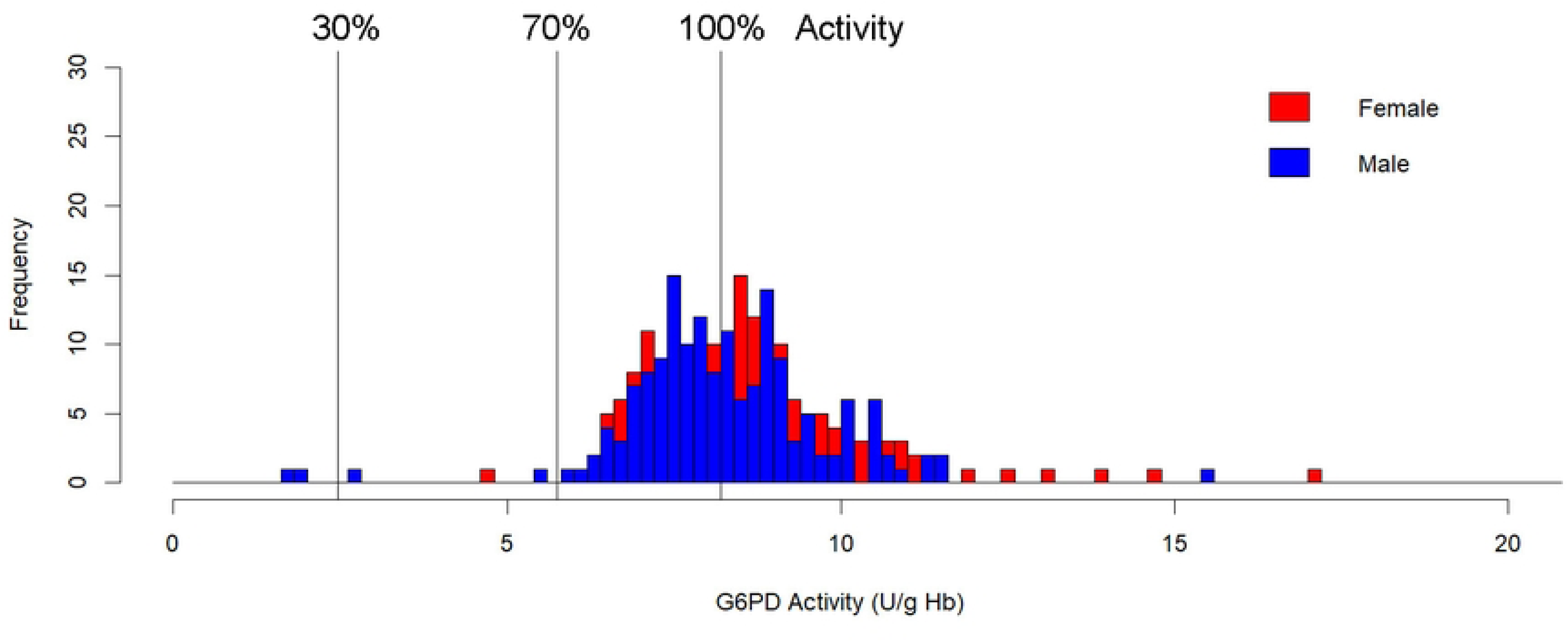
G6PD activity distribution curves in both males and females across schoolchildren in Tanjung Leidong.

Subjects with <30% of G6PD activity are considered deficient, 30-70% as intermediate, and >70% as normal.

### Association of body mass index with anemia status and RBC disorders

Comparison analysis with hemoglobin levels was done to observe any association between nutritional status and present conditions like varying degrees of anemia. Fig 4 shows a trend where the proportion of patients with anemia and RBC disorders was highest among underweight individuals and decreased progressively across normal, overweight, and obese BMI categories. Statistical analysis showed no significant association between BMI and anemia (p = 0.274). Meanwhile, there was a weak negative correlation between BMI and RBC disorders (p = 0.039).

**Fig 4.**
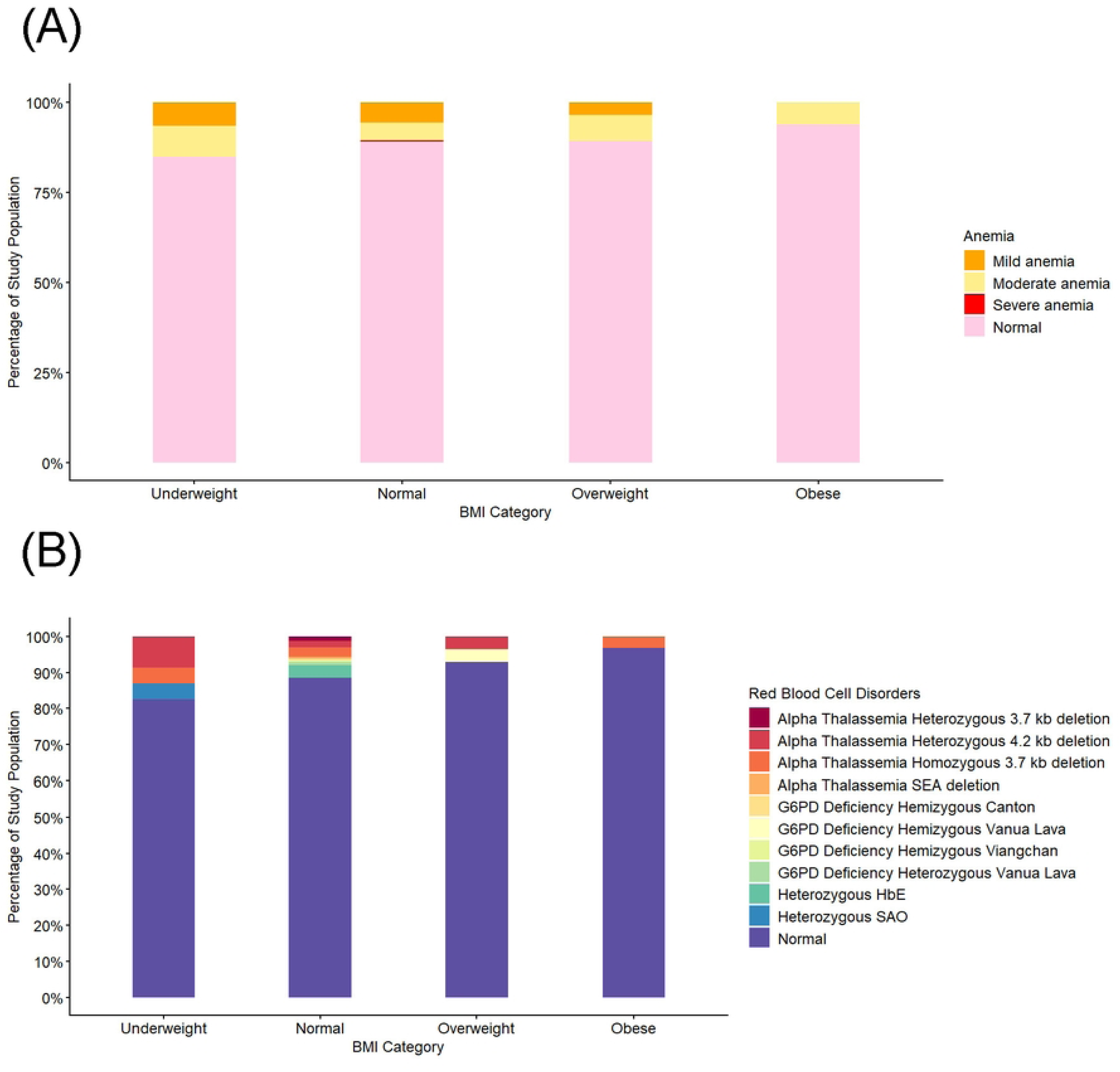
Comparison of BMI category against (A) anemia status and (B) RBC disorders.

### Hemoglobin levels in subjects with RBC disorders

Subjects with RBC disorders were compared based on their hemoglobin levels, which indicated RBC quantities. As seen in Table 3, there was no significant difference between hemoglobin level in normal and samples with RBC disorders. It was observed that heterozygous one-gene deletion had lower levels of hemoglobin than the homozygous state. The same could be said for the SEA deletion. However, hemoglobin levels for heterozygous SAO and HbE were slightly lower than normal samples, though all of this was statistically significant. Generally, the male hemoglobin levels overall outweigh the females.

**Table 3.** Hemoglobin level comparison in RBC disorders found in Tanjung Leidong.

| Variable | Hemoglobin (g/dL)<sup>a</sup> |  |
| --- | --- | --- |
|  | Female | Male |
| **G6PD** |  |  |
| Deficient | - | 15.43 (0.81) |
| Intermediate | 13.25 (1.34) | 10.60 (NA) |
| Normal | 13.05 (1.22) | 13.96 (1.93) |
| p-value | 0.8<sup>c</sup> | 0.065<sup>b</sup> |
| **SAO** |  |  |
| Heterozygous | 12.95 (1.91) | 16.90 (NA) |
| Normal | 13.06 (1.21) | 13.95 (1.93) |
| p-value<sup>c</sup> | >0.9 | 0.13 |
| **HbE** |  |  |
| Heterozygous | 12.54 (1.04) | 12.50 (NA) |
| Normal | 13.08 (1.22) | 13.98 (1.94) |
| p-value<sup>c</sup> | 0.2 | 0.3 |
| **α-Thalassemia One-Gene Deletion** |  |  |
| Homozygous 3.7 kb deletion | 12.98 (0.80) | 15.73 (2.29) |
| Heterozygous 3.7 kb deletion | 12.90 (0.99) | 12.00 (NA) |
| Heterozygous 4.2 kb deletion | 12.95 (1.10) | 12.50 (0.98) |
| Normal | 13.06 (1.24) | 13.99 (1.93) |
| p-value<sup>b</sup> | >0.9 | 0.054 |
| **α-Thalassemia Two-Gene Deletion** |  |  |
| Heterozygous --<sup>SEA</sup> | 9.50 (NA) | 0 (0.00) |
| Normal | 13.07 (1.19) | 13.97 (1.94) |
| p-value<sup>c</sup> | 0.091 | - |

<sup>a</sup>Mean (SD)
<sup>b</sup>Kruskal-Wallis rank sum test
<sup>c</sup>Wilcoxon rank sum test

### Measurement of hemoglobin using different devices

Hemoglobin detection using hematology analyzer remains the gold standard for hemoglobin measurement, however in this study, HemoCue will be used as the gold standard against the STANDARD G6PD Analyzer for hemoglobin measurement. As shown in Fig 5, a Bland-Altman plot was generated to compare the two detection methods. A higher total number of dots near the mean line indicated comparable results between the two devices. A mean of 0.709 means that HemoCue measures the hemoglobin at 0.709 higher or lower than the STANDARD G6PD Analyzer, indicating that the two methods are not interchangeable.

**Fig 5.**
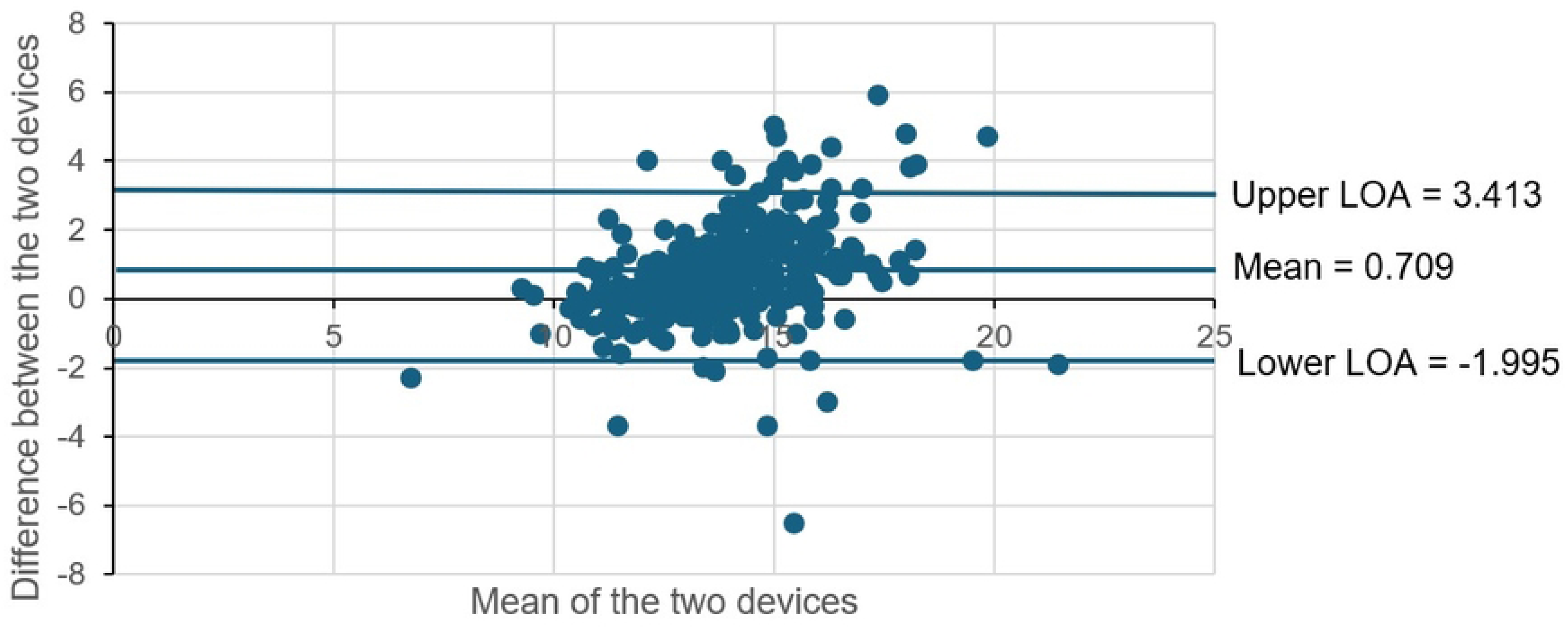
Comparison of hemoglobin value measured by HemoCue and STANDARD G6PD Analyzer using the Bland-Altman plot. LOA = limit of agreement.

## Discussion

Our study reveals several RBC disorders found in schoolchildren in Tanjung Leidong. Even though malaria is negative in all the cohorts, asymptomatic malaria infections cannot be ruled out, particularly since our study population consisted primarily of non-febrile patients. Asymptomatic malaria contributes to higher detection of RBC disorders as it causes subclinical hemolysis and anemia, which unmasks mild traits of disorders [29, 30]. Unfortunately, we did not employ PCR in this study to detect malaria parasites.

The proportion of SAO in this study is low (<1%). This might contribute to the low malaria cases in Tanjung Leidong, as some studies concluded that SAO protects against cerebral malaria [31]. Another study in Nias District in 2007 recorded that SAO prevalence was only 1% even though malaria positivity was around 8% [32]. The mechanism of said protection remains unclear, albeit it has been proposed that the rigid RBC membrane interferes with merozoite invasion and prevent parasitic development in RBCs [33]. Aikawa (1988) proposed alterations that could affect binding to the RBC membrane. Some protection also come in the form of redistribution of infected RBCs away from the brain [35]. HbE also has a similarly low proportion under 1%, which is comparable to a study by Nuryanti et al. (2015), in which they recorded a prevalence of 0.7% (4/560) in Medan, North Sumatra. Similar to SAO, HbE has been noted to protect against malaria, albeit the source is indeterminate [2]. HbE-carrying erythrocytes may reduce susceptibility to malaria by impairing parasite invasion and growth and enhancing immune clearance of infected erythrocytes [8, 37]. Although severe clinical symptoms for SAO and HbE do not manifest in their heterozygous state, some precautions should be taken as less deformable RBCs could be prone to getting caught in the microvasculature of the spleen, thus getting eliminated by the spleen in the case of malaria infection [2].

Previous G6PD deficiency survey in Tanjung Leidong had shown similar prevalence data with our study at 2.6% [28] with G6PD Mahidol (c.487 G > A) as the dominant variant, unlike ours, which were Vanua Lava (c.383 T > C) and Viangchan (c.871 G > A). A study by Wisnumurti et al. (2019) stated that the most common variants found in Deutromalay populations are Viangchan, Canton (c.1376 G > T), and Vanua Lava. As the predominant ethnicities in our study consisted of Batak, Javanese, and Chinese, our findings in this study aligned with regional patterns where those variants predominate.

Anemia proportion of early adolescent women almost doubles that of men, which is consistent with previous studies, as women lose more blood during menstruation and need more iron in their diet compared to men [39]. Even though that is the case, most α-thalassemia mutations had normal hemoglobin levels. Moreover, anemia could also be caused by nutritional and iron status as well as genetic factors, and based on this study’s observation, nutritional factors may affect anemia proportion in Tanjung Leidong (Fig 4A). However, determination of such a conclusion requires more complete blood screening, such as mean corpuscular volume value, HbA2, HbF, and iron deficiency analyses [14]. Carriers of the thalassemia trait might not manifest symptoms or exhibit mild symptoms and thus, their hemoglobin levels would remain normal [40] as shown in Table 3 where homozygous and heterozygous conditions do not differ from normal. The proportions of α-thalassemia are the highest among other RBC disorders, about 6%. This would explain the fact that malaria incidence remains low despite low proportions of SAO, HbE, and G6PD deficiency. It was investigated that one-gene deletion was associated with reductions in admission rate of patients with malaria and severe malaria, indicating some protection provided to the host. However, it does not protect against asymptomatic malaria [41]. Meanwhile, one child was found to have α-thalassemia --^SEA^ mutation, which is the most prominent in Southeast Asia. As a two-gene deletion would have caused a more significant α-globin gene loss, a reduced amount of hemoglobin levels (often below 10 g/dL) is expected [42] (Table 3).

Underweight children show higher proportions of RBC disorders and anemia status compared to the other categories (Fig 4A & Fig **4**B), which, together with nutritional deficiencies, may exacerbate erythropoiesis and hemoglobin synthesis, thus worsening the anemia and increasing the susceptibility to infections and stunting [43–45]. Although asymptomatic, when compounded with factors such as underweight, malnutrition worsens α-globin levels, which also affects erythropoiesis [46]. However, we also see mild-moderate anemia in children who are overweight and obese with RBC disorders.

Taken together, these results highlight that low BMI and RBC disorders do not act independently but instead interact in a manner that aggravates hematological outcomes, underscoring the need for integrated nutritional and genetic interventions.

Based on the Bland-Altman plot (Fig 5), the agreement between hemoglobin measurements obtained using the HemoCue and the STANDARD G6PD Analyzer was inadequate, indicating that the STANDARD G6PD Analyzer is not yet sufficient to replace the HemoCue for accurate hemoglobin measurement. While most dots shown in the plot were centered toward the mean, several outliers gave rise to a wide difference in limits of agreement of 5.408 g/dL (-1.995 to 3.413). A similar study had similar results, where there was a 2.3–3.1 g/dL difference in measurement between the devices [47]. Based on a study by Adam et al. (2012), a clinically acceptable difference between limits of agreement during hemoglobin measurement is roughly 1 g/dL, from which this study’s comparison greatly exceeds. Furthermore, a difference of 0.709 g/dL in hemoglobin level would influence G6PD activity and deficiency determination. The accuracy of hemoglobin measurement is critical in high-dose primaquine prescription to prevent hemolysis of G6PD-deficient patients.

Some limitations are observed during this study. Although representative, the small sample size provides limited statistical power and generalization. The samples collected were from clinically healthy subjects and were not reconfirmed for asymptomatic malaria using PCR which could act as a contributing factor to anemia and stunting down the road. Lack of blood microscopy prevented a more thorough evaluation of the RBC morphologies. We also did not screen for iron deficiency to differentiate the contributing factor of anemia in this study. The absence of a hematological analyzer as the gold standard for hemoglobin measurement limited our ability to determine accurate hemoglobin levels and prevented comprehensive hematological profiling.

Detection of RBC disorders is vital in alleviating anemia symptoms that may arise due to hemolysis, whether due to infections or, in the case of G6PD deficiency, oxidative triggers. This study encourages government-led screening to enable early identification of RBC disorders, particularly in malaria-endemic areas. Systematic screening of RBC disorders facilitates the detection of asymptomatic carriers and provides education for these carriers to reduce symptom manifestation. Furthermore, comprehensive screening programs can support genetic counselling efforts, helping families to prevent the inheritance of harmful variants to future generations.

## Conclusions

This study is a first step in profiling RBC disorders in the malaria-endemic region of Tanjung Leidong, providing important insights into the distribution of RBC disorders in this area. Some RBC disorders are detected in schoolchildren, which indicates how important genetic screenings are to provide awareness for the manifestation of symptoms and triggers. The findings in this study suggest that STANDARD G6PD Analyzer by SD Biosensor should not be used for hemoglobin measurement and the measurement should still be done by the gold standard or HemoCue. Although this study has limitations, the results show that the mapping of RBC disorders is in line with previous studies and can serve as a basis for further research and health policy development, particularly in malaria elimination and stunting in children

## Data Availability

Data available on request from authors.

## Acknowledgements

We would like to thank all research participants and teachers in the D. I. Panjaitan Foundation, Tanjung Leidong, as well as the Labuhanbatu Utara District Health Office and local health workers. Our appreciation also goes to the laboratory team for data analysis and testing for RBC disorders, the field staff, as well as to all those who have contributed to this study.

